# Trends in Fracture-Related Hospitalizations and Mortality in Brazil, 2015–2024 Bone Fracture Trends in Brazil

**DOI:** 10.64898/2025.12.23.25342914

**Authors:** Palloma Porto Almeida, Danielle Cabral Bonfim

## Abstract

**Background:** Bone fractures represent a growing public health concern worldwide, yet national epidemiological assessments remain limited in Brazil. Understanding temporal trends, demographic disparities, and geographic heterogeneity is essential to guide prevention, resource allocation, and trauma-care planning.

**Objective:** To characterize the epidemiological profile of bone-fracture–related hospitalizations and mortality in Brazil between 2015 and 2024, analyzing trends by sex, age, fracture type, and geographic and ethnic distribution.

**Methods:** An epidemiological, observational, descriptive, and population-based ecological study was conducted using SIH/SUS and IBGE data. Hospitalization rates, case fatality rates (CFR), relative risks, odds ratios, and Years of Life Lost (YLL) were calculated. Temporal trends were evaluated using Annual Percent Change (APC).

**Results:** Other limb fractures were the most frequent injuries, while femur fractures showed the highest lethality. Men had nearly twice the hospitalization rate of women, driven by high-energy trauma in adults aged 20–59 years, whereas women experienced a sharp increase in femur-fracture admissions at older ages. Skull, facial, and thorax/pelvis fractures contributed disproportionately to premature mortality. Marked geographic and ethnic disparities were observed, with higher burdens in the North/Northeast and predominance among Brown and Indigenous populations.

**Conclusion:** Fracture-related hospitalizations in Brazil have increased consistently, with distinct epidemiological patterns across demographic and regional groups. These findings highlight the need for targeted prevention and improved trauma-care strategies.

**Key Messages:**

- This study investigated national temporal trends and demographic, geographic, and ethnic disparities in bone-fracture–related hospitalizations and mortality in Brazil from 2015 to 2024.
- We found a sustained increase in fracture-related hospitalizations, with femur fractures showing the greatest lethality and fractures of other limb bones accounting for the highest volume, and marked disparities by sex, age, region, and ethnicity.
- These findings are important because they provide the first comprehensive national overview of fracture burden in Brazil and offer evidence to inform targeted prevention policies, equitable resource allocation, and trauma-care planning.

## Introduction

Bone fractures represent a major public health concern worldwide and play an important role in morbidity across all age groups. From high-energy trauma in young adults to fragility fractures in older populations, these injuries are associated with substantial mortality, long-term disability, and significant reductions in quality of life (Cauley 2013; Wu *et al*. 2021). Their impact extends beyond clinical complications, encompassing considerable direct costs related to hospitalization, surgical procedures, and rehabilitation, as well as indirect costs associated with loss of productivity, long-term caregiving demands, and reduced functional independence (Mensor *et al*. 2021; Sarmento *et al*. 2022).

In Brazil, understanding the epidemiology of fractures is essential for guiding public health planning, improving resource allocation, and informing preventive strategies. The burden of these injuries varies according to demographic and social factors, including sex, age, and ethnic–racial background, mirroring global patterns of vulnerability (Albergaria *et al*. 2022). Despite their relevance, systematized national data on hospitalization profiles, fracture-related mortality, and economic impact remain limited. However, the availability of robust population-based databases, particularly the National Hospital Information System (SIH/SUS) provides a valuable opportunity to characterize the magnitude and distribution of fracture-related hospitalizations at a national level.

Leveraging these databases enables the identification of temporal trends, regional disparities, sex-specific patterns, and risks associated with different fracture types, contributing to a more comprehensive understanding of the clinical and socioeconomic burden of fractures in the country. Therefore, the aim of this study is to characterize the epidemiological profile of bone fractures in Brazil from 2015 to 2024, examining trends in hospital admissions and mortality stratified by sex, age, year of occurrence, state, and type of fracture.

## Material and Methods

### Data collection

This is an epidemiological, observational, descriptive, and population-based ecological study. The hospitalization and death data related to bone fractures were obtained from public domain sources, provided by the Hospital Information System of the Unified Health System (SIH/SUS) through the health information platform Tabnet via the electronic portal of the Department of Informatics of the Unified Health System (DATASUS). The dataset was extracted between August 18th and August 21st, 2025, and stratified by sex, age, ethnicity, year, and state of hospitalization, referring to the period from 2015 to 2024. Life expectancy data were obtained from the Brazilian Institute of Geography and Statistics (IBGE), with the most recent available data from 2023, stratified by sex and age. Annual estimates of the resident population by age group in Brazil were obtained from DATASUS/IBGE for the same period (2015–2024).

### Calculation of hospitalization rates

Hospitalization rates per 100,000 inhabitants were calculated as follows:

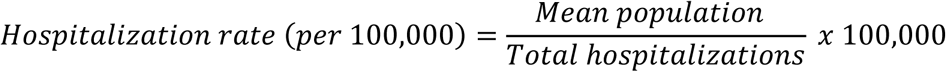

This approach allowed for comparison of fracture burden between men and women, adjusted for population size.

### Calculation of Years of Life Lost (YLL)

This approach is analogous to the person-years of life lost (PYLL), although here it was applied specifically to fracture mortality rather than the total population. Data from SIH/SUS were displayed according to the Brazilian Ministry of Health categories: <1 year, 1–4 years, 5–9 years, 10–14 years, 15–19 years, 20–29 years, 30–39 years, 40–49 years, 50–59 years, 60–69 years, 70–79 years, and ≥80 years.

The total deaths in each age group were evenly distributed across all ages within the group to calculate deaths per age. Years of Life Lost (YLL) were calculated for fracture-related deaths using individual-level mortality data stratified by sex and age group. First, age groups were expanded into exact ages for each death, creating a dataset where each row represented a single death with the corresponding age 𝑎_𝑖_ and sex 𝑠_𝑖_.

The life expectancy 𝐿𝐸(𝑎_𝑖_, 𝑠_𝑖_) for each individual was obtained from national life tables. For each death, YLL was defined as:

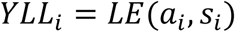

The total YLL for each fracture category 𝑐 and sex 𝑠 was calculated as the sum over all deaths:

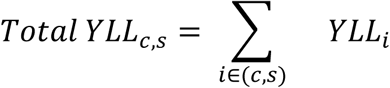

The mean YLL per death was computed as:

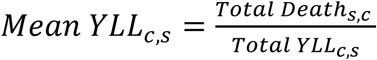

Finally, the proportion of total YLL attributable to each fracture category was calculated as:

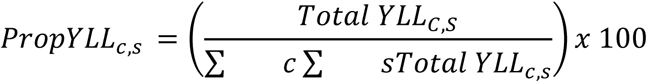

This approach provides both the absolute burden in years of life lost and the average YLL per death, allowing interpretation in terms of the typical years of life lost per fatal fracture, as well as the relative contribution of each fracture type to the overall YLL.

### Case Fatality Rate (CFR)

Case Fatality Rates (CFR) was calculated to estimate the lethality associated with each fracture type. CFR (%) were calculated as the proportion of in-hospital deaths among all hospitalizations for each fracture category, sex, and year:

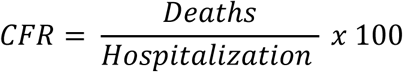

To represent overall lethality by fracture type, data were aggregated across the entire study period (2015–2024), and an aggregate CFR (%) was computed for each fracture category as:

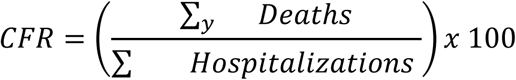

### Statistics

Trends in fracture-related hospitalizations from 2015 to 2024 were analyzed using the Annual Percent Change (APC), calculated through a log-linear regression model. The APC and its 95% confidence interval were estimated to quantify the average yearly change in hospitalization rates. Hospitalizations were analyzed by age group, and hospitalization rates per 100 000 inhabitants were calculated using the mean population of each age group.

The chi-square test of independence was applied to evaluate the association between sex and fracture type. The test’s applicability conditions were verified: no cell in the contingency table had an expected frequency less than 1, and 100% of the cells had expected frequencies ≥ 5, ensuring the validity of the test. Sex-specific odds ratios with 95% confidence intervals were calculated for both hospitalization and mortality, allowing comparison of the relative risk of hospitalization and death between women and men for each fracture type.

To compare YLL between sexes for each fracture type, individual-level observations were generated from absolute mortality data. Each death reported in the hospital dataset was expanded into a separate observation with the corresponding age of death assigned according to the midpoint of the reported age group. The YLL for each death was then calculated as the remaining life expectancy at the age of death, based on national life tables from IBGE. This procedure allowed us to treat the data as if each row represented an artificial individual’s death. Then a two-sample t-test was applied to compare the mean YLL between males and females for each fracture type.

Differences in the national distribution of fracture-related hospitalizations across ethnic groups were first assessed using a chi-square goodness-of-fit test. After detecting a significant overall difference, pairwise chi-square tests for equality of proportions were conducted. Bonferroni correction was applied to adjust for multiple comparisons.

In all equations, subscripts were used to denote specific dimensions of the dataset:

● 𝑐 represents the fracture category, according to ICD-10 classification
● 𝑠 denotes sex (male or female);
● 𝑦 indicates the year of observation (from 2015 to 2024); and
● 𝑖 refers to individual deaths within each combination of 𝑐, 𝑠, and 𝑦.

All Statistical analyses were performed using R software, significance was defined as P < 0.05.

## Results

### Trends in Hospitalizations between 2015 and 2024

The temporal analysis (2015–2024) revealed consistent upward trends in hospitalization rates across all fracture categories (Figure 1. Femur fractures showed a consistent linear increase over time, albeit at lower absolute volumes. In contrast, fractures of other limb bones exhibited both the highest baseline volume and the strongest linear growth, reinforcing their role as the primary contributor to fracture-related morbidity nationwide. A distinct deviation was observed for skull and facial fractures, which experienced a transient decline in 2020. This drop aligns with the period of restricted mobility, reduced traffic flow, and lower rates of interpersonal violence associated with COVID-19 mitigation measures. After 2021, the hospitalization rates for this fracture type resumed their upward trend, consistent with the patterns of the pre-pandemic years.

**Figure 1.**
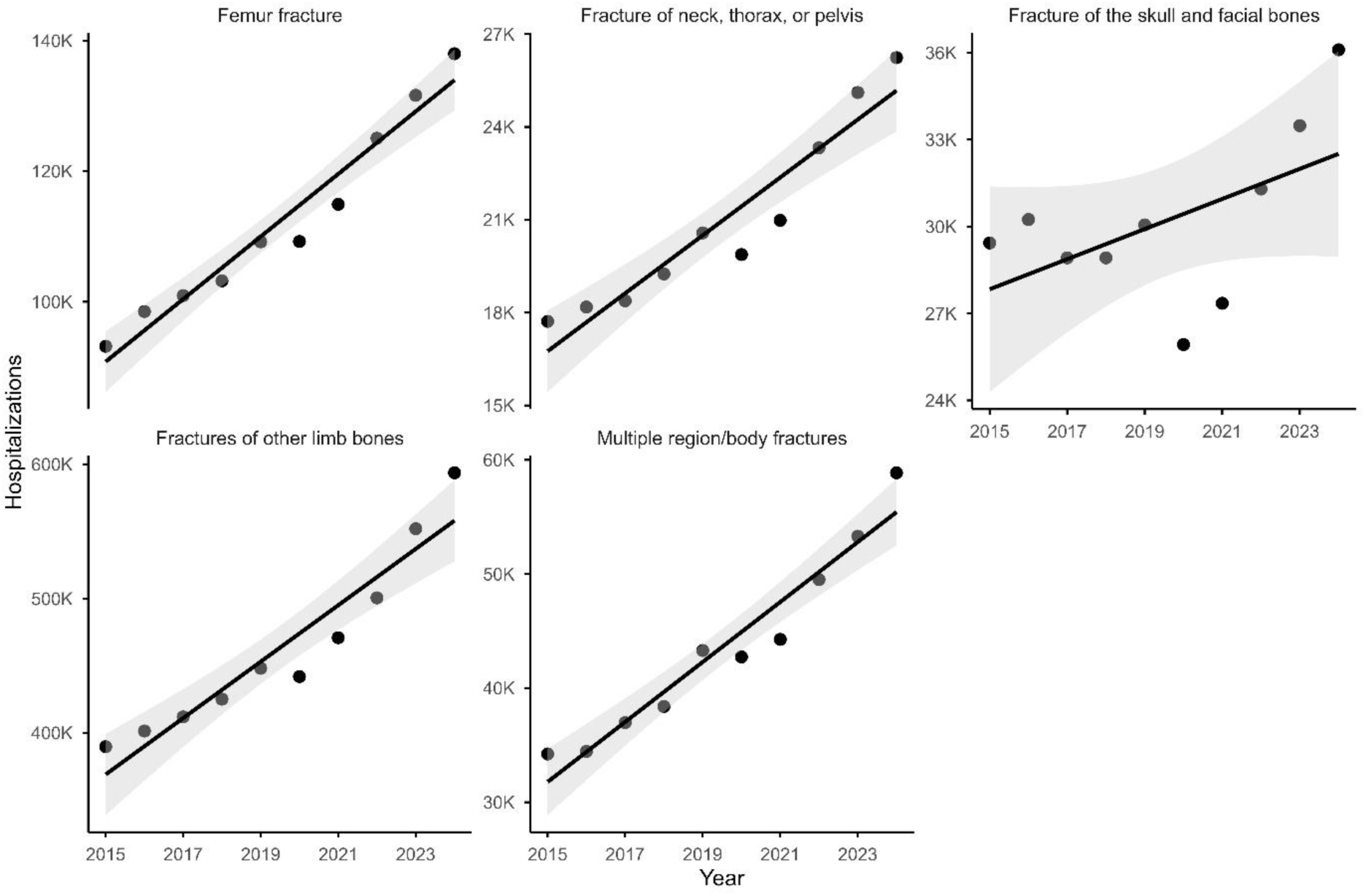
Annual trends in fracture-related hospitalizations in Brazil from 2015 to 2024, by fracture type. Data points represent observed annual hospitalization counts for each fracture category, and the solid line indicates the fitted linear trend with its 95% confidence interval. All values are shown on the original (non-logarithmic) scale.

Analysis of hospitalization trends by fracture type, adjusted for population size, revealed variable annual percent changes (APC) across the different fracture categories. Multiple region/body fractures exhibited the highest increasing trend of hospitalizations, with an APC of 6.13% per year from 2015 to 2024, followed by fractures of other limb bones, displaying an APC of 4.51%. Femur fractures and fractures of the neck, thorax, or pelvis increased at a similar rate. All analyses were statistically significant, except for fractures of the skull and facial bones that showed a non-significant trend, indicating a relatively stable rate over the study period (1.61%).

These findings highlight differential trends in fracture-related hospitalizations, with most categories showing a statistically significant increase over time (Table 1).

### Fatality

The analysis of fracture-related case fatalities (CF) reveals significant variation by fracture type. Femur fractures presented the highest fatality rate (3.1%), followed by fractures of the neck, thorax, or pelvis (2.6%), multiple region fractures (1.8%), skull and facial fractures (0.8%), and other limb fractures (<0.5%). These findings highlight the importance of identifying fractures with higher mortality risk to guide public health policies, prioritize preventive strategies, such as fall prevention and traffic safety, and optimize trauma care and resource allocation for the most vulnerable patient groups.

Analysis of CF by fracture type between 2015 and 2024 revealed marked heterogeneity in severity. Femur fractures consistently exhibited the highest lethality, with annual CFs ranging from 2.9% to 3.5%, and a notable peak in 2021 (3.46%), underscoring their clinical impact on mortality. Fractures of the neck, thorax, or pelvis also showed elevated CFs (2.0–2.8%), with a slight decline around 2020 followed by recovery in subsequent years. Multiple region fractures maintained intermediate lethality, fluctuating between 1.6% and 2.1%, while skull and facial fractures had lower CFs (<0.8%), and fractures of other limb bones remained the least fatal (<0.2%) despite being the most frequent (Figure 2). Importantly, temporal trends suggest relative stability across most categories, with transient variations in 2020 likely reflecting healthcare disruptions during the COVID-19 pandemic.

**Figure 2.**
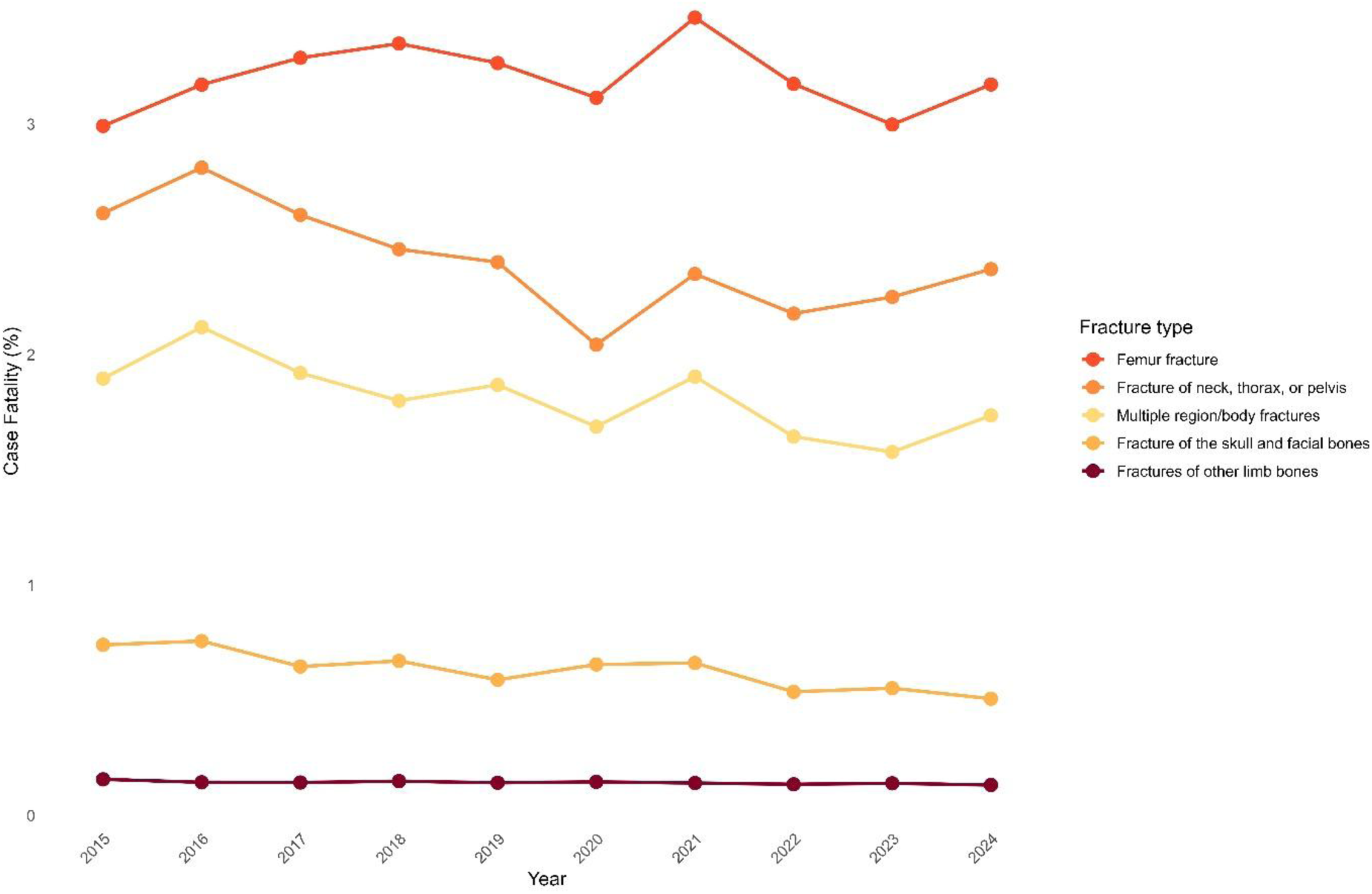
Annual case fatality rates (%) for fracture-related hospitalizations in Brazil, 2015–2024, by fracture type. Case fatality rate was defined as the percentage of in-hospital deaths among all hospitalizations for each fracture category. Values represent annual percentages based on SIH/SUS records.

### Sex differences in hospitalization rates

In the analysis of hospitalizations among men and women from 2015 to 2024, adjusted for population size, men exhibited a higher cumulative rate of 4600 per 100 000 inhabitants, approximately twice that of females (2265 per 100 000 inhabitants), indicating a greater overall risk of fracture-related hospitalizations among men. A chi-squared test was applied to evaluate the association between sex and fracture type. The analysis revealed a highly significant result (χ² = 57,092, df = 4, p < 2.2e-16), indicating that the distribution of hospitalizations due to fractures is not independent of sex.

Analyzing the hospitalizations by fracture type, women presented a lower absolute number of hospitalizations due to femur fractures compared with men. However, within the overall distribution of fracture types among females, femur fractures were the second leading cause of admissions, accounting for 16.5% of all female fracture-related hospitalizations. Notably, women had almost twice the odds of experiencing a femur fracture, which is clinically relevant given the higher fatality rate observed in females (6.86%) compared with males (2.83%). Fractures of other limb bones represented the major cause of hospitalizations in both sexes, with similar proportional contributions (71.6% in women and 73.4% in men); despite their high frequency, these fractures showed very low fatality rates (0.14–0.16%). In contrast, sex-specific patterns became more pronounced for fractures associated with high-energy trauma: men displayed considerably higher odds of sustaining skull and facial fractures (OR 0.48) and fractures of the neck, thorax, or pelvis (OR 0.80). These injuries were markedly more frequent in males (256 554 skull/facial fractures vs. 56,994 in females), suggesting differences in exposure to occupational hazards, risk-taking behaviors, or external trauma mechanisms. Altogether, these findings reveal distinct epidemiological profiles of fracture-related hospitalizations and outcomes between men and women (Table 2).

When analyzing hospitalizations by sex and age group, fractures of other limb bones accounted for the highest number of admissions among men, totaling 2 871 835 hospitalizations between 20 and 59 years of age. All fractures, except for femur fractures, followed the same pattern related to number of admissions in male gender. The age group with the greatest concentration of cases was 20–29 years, with 754,956 admissions, forming the first peak of a clear bimodal pattern in males that reflects high-energy trauma in young adulthood and a secondary increase after 50 years. In contrast, women exhibited a markedly different distribution: femur fractures were the leading cause of hospitalization in older adults, with 242 702 admissions in those aged 80 years or older, followed by fractures of other limb bones among women aged 60–69 years (236 501 hospitalizations) (Supplementary file 1). The heatmap highlights these divergent epidemiological patterns, young and middle-aged men concentrated in trauma-related fractures, particularly of other limb bones, whereas fracture-related admissions among women rise sharply only in later life, consistent with age-related bone fragility. Skull, facial, and thorax/pelvis fractures were also more prominent among men aged 20–49 years, further reinforcing the sex-specific mechanisms underlying fracture risk (Figure 3).

**Figure 3.**
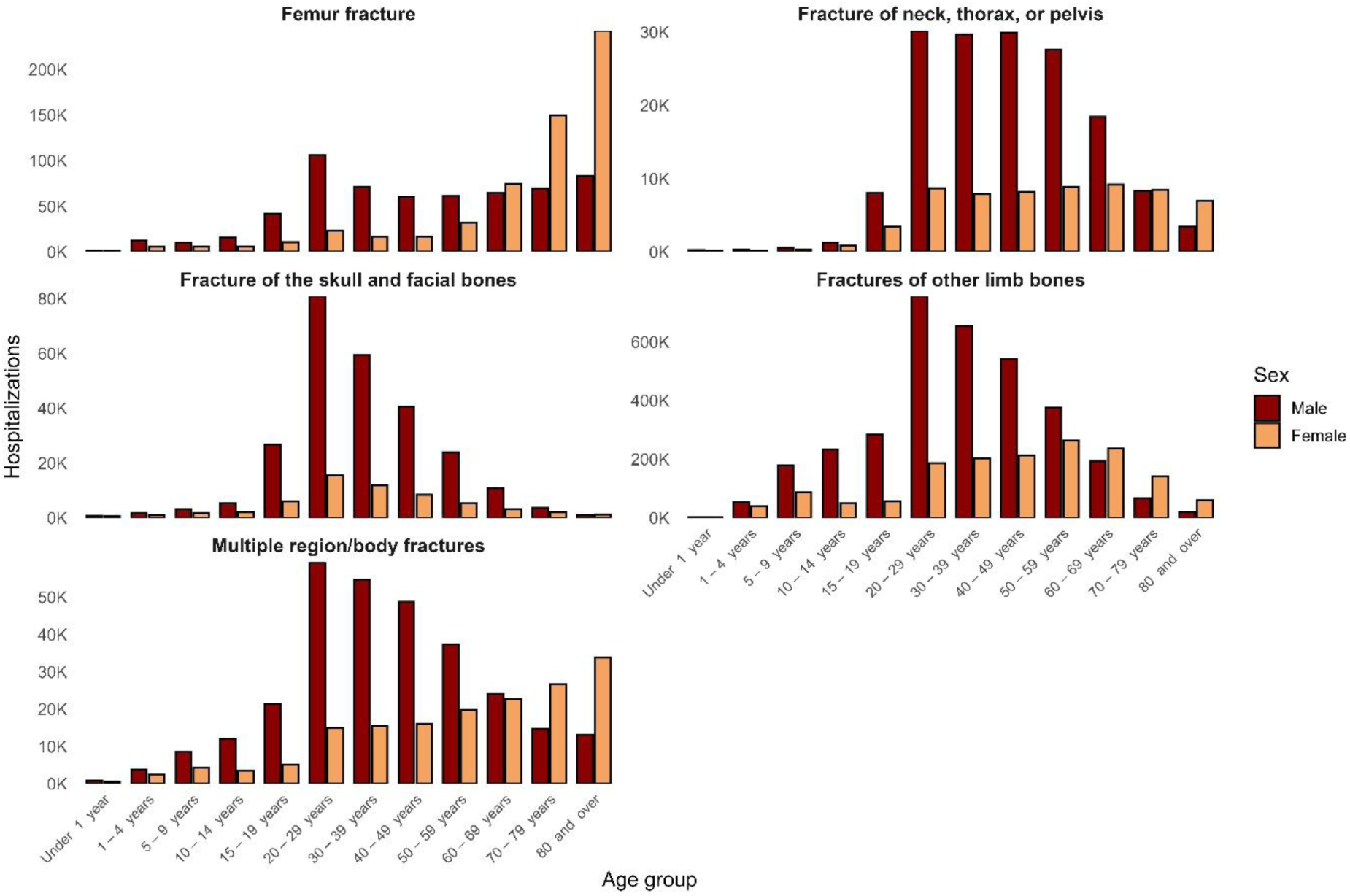
Distribution of fracture-related hospitalizations in Brazil from 2015 to 2024 by sex, age group, and fracture type. Stacked panel bar charts display the absolute number of hospitalizations for each fracture category across age groups, stratified by sex. Bars represent annual counts aggregated over the study period, allowing comparison of age- and sex-specific hospitalization patterns within each fracture type.

### Years of Life Lost by Fracture Type

Analysis of YLL due to fracture-related deaths revealed substantial differences by fracture type and sex. Femur fractures accounted for the largest share of total YLL, representing 25.8% in females and 22.3% in males, with mean years of life lost of 10 and 14 years, respectively, consistent with mortality occurring predominantly at older ages. In contrast, fractures of the neck, thorax, or pelvis contributed proportionally less to total YLL (2.97% in females and 11.3% in males), but exhibited higher mean YLL per death (23 and 27 years), indicating deaths at earlier ages, particularly among men. Skull and facial fractures showed the smallest contribution to overall YLL (0.92% in females and 5.6% in males) yet had the highest mean YLL (32 and 34 years), highlighting their disproportionate impact on premature mortality. Fractures of other limb bones accounted for 4.6% of total YLL in females and 13.4% in males, with mean YLL of 19 and 28 years, while multiple-region fractures contributed 5.3% and 7.8%, with 11 and 19 mean years lost, respectively (Table 3).

Across all fracture categories, males experienced systematically higher mean YLL per death, underscoring a greater burden of premature mortality. Collectively, these patterns suggest that although femur fractures dominate the total YLL burden due to their frequency and fatality in older adults, high-energy trauma fractures, such as skull/facial and thorax/pelvis injuries, carry the greatest risk of premature death, particularly among men. All sex comparisons in mean YLL were statistically significant (P < 0.001).

### Geographical distribution of hospitalizations

Population-adjusted hospitalization rates for bone fractures varied widely across Brazilian states (Figure 4). The highest rates were concentrated in Rondônia (4745 per 100 00), Piauí (4402), and Mato Grosso (4200), followed by intermediate values in the South and Southeast (2500–3500 per 100 000). In contrast, Amazonas and Acre recorded the lowest hospitalization rates, below 2000 per 100 000 inhabitants (Supplementary file 2). These regional differences may reflect variations in healthcare access, trauma incidence, reporting systems, and demographic structure, rather than differences in age composition. Overall, the data reveal a spatially heterogeneous distribution of fracture-related hospitalizations.

**Figure 4.**
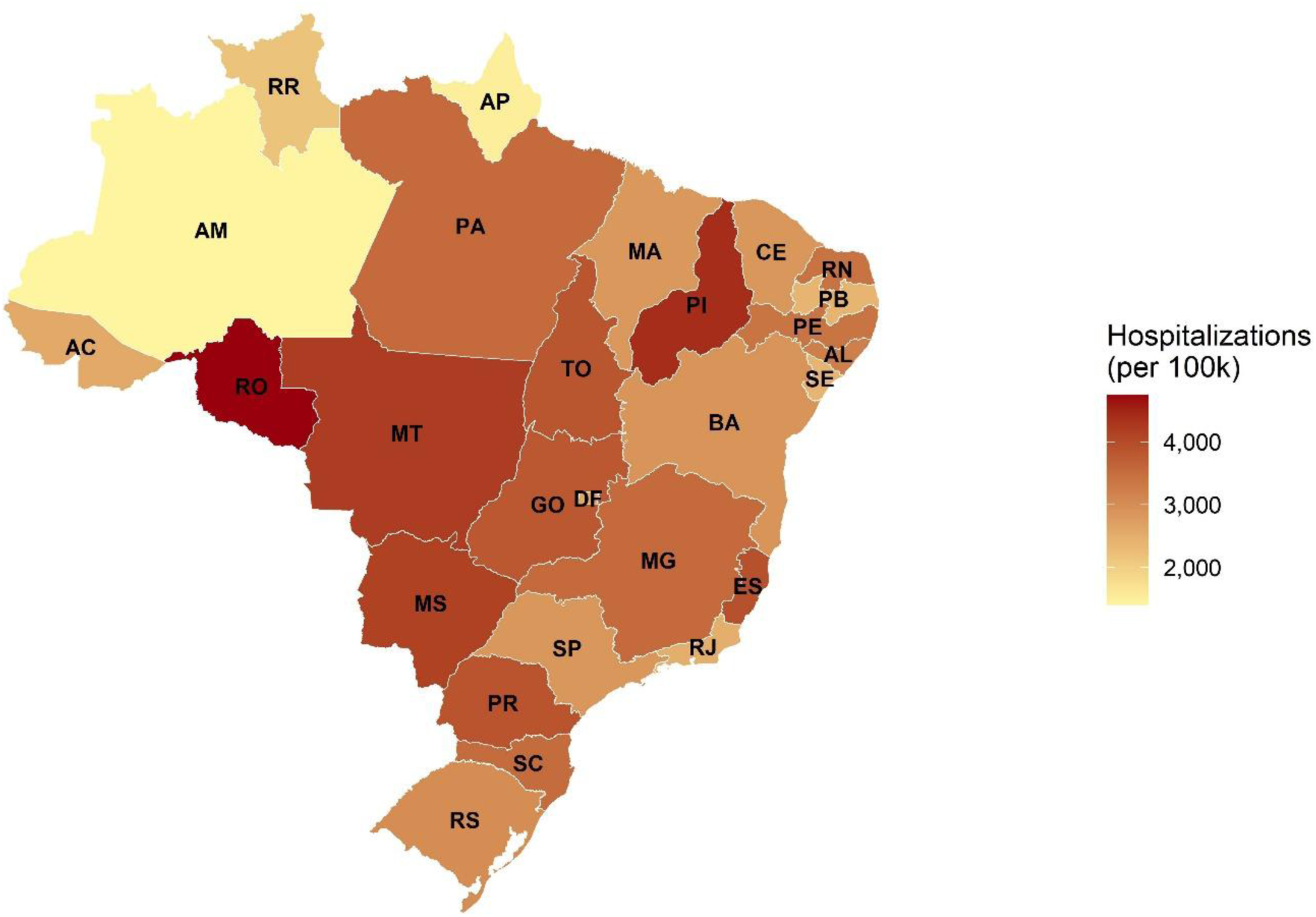
Population-adjusted hospitalization rates for bone fractures across Brazilian states, 2015–2024. Choropleth map showing hospitalization rates per 100,000 inhabitants for all fracture types combined. Rates were calculated using SIH/SUS hospitalization records and population estimates from IBGE. Colour intensity represents the magnitude of the rate in each state.

### Ethnic distribution of hospitalizations

Related to the ethnicity profile of the hospitalized patients, the majority were Brown (Mixed), corresponding to 55.99% of all cases, followed by White ethnicity (37.48%). All pairwise differences between ethnic groups were statistically significant (Bonferroni-adjusted P < 0.05), indicating that the distribution of fracture-related hospitalizations differs meaningfully across ethnic groups and that each ethnicity contributes a distinct proportion to the overall hospitalization burden (Figure 5).

**Figure 5.**
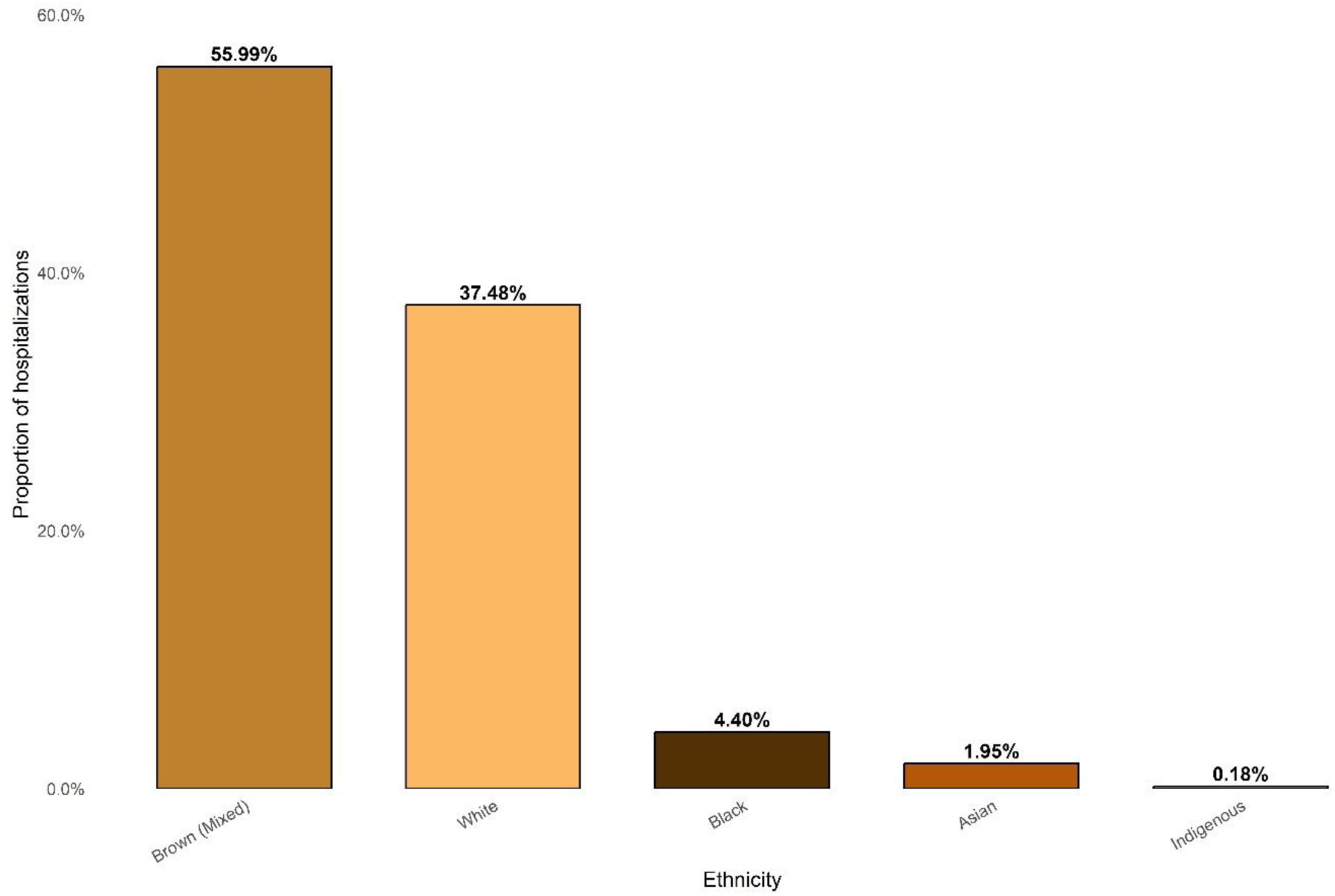
Proportion of fracture-related hospitalizations in Brazil, 2015–2024, by ethnicity. Proportions represent the distribution of SIH/SUS-recorded hospitalizations across ethnic groups. Pairwise differences between ethnic groups were assessed following a significant chi-square test, with P-values adjusted using the Bonferroni correction (P < 0.05).

Regarding the place of hospitalizations, the number was highest in Minas Gerais for Asian and Brown (Mixed) individuals, and in São Paulo for Black and White individuals (in absolute numbers). However, the proportional analysis reveals a different pattern. Some states with smaller absolute numbers contributed a larger share of hospitalizations within specific ethnic groups, highlighting regional disparities in hospitalization rates relative to the size of each ethnic population. Brown (Mixed) individuals were the predominant group among hospitalizations across most states. Their representation exceeded 50% in nearly all regions, with particularly high proportions in states such as Pará (94.2%), Pernambuco (92.9%), Tocantins (91.1%), Piauí (89.7%), and Bahia (87.1%). The only exceptions were Paraná, Rio Grande do Sul, Santa Catarina, and São Paulo, where White individuals were the majority, accounting for 76.0%, 88.8%, 89.2%, and 60.4% of hospitalizations, respectively (Figure 6).

**Figure 6.**
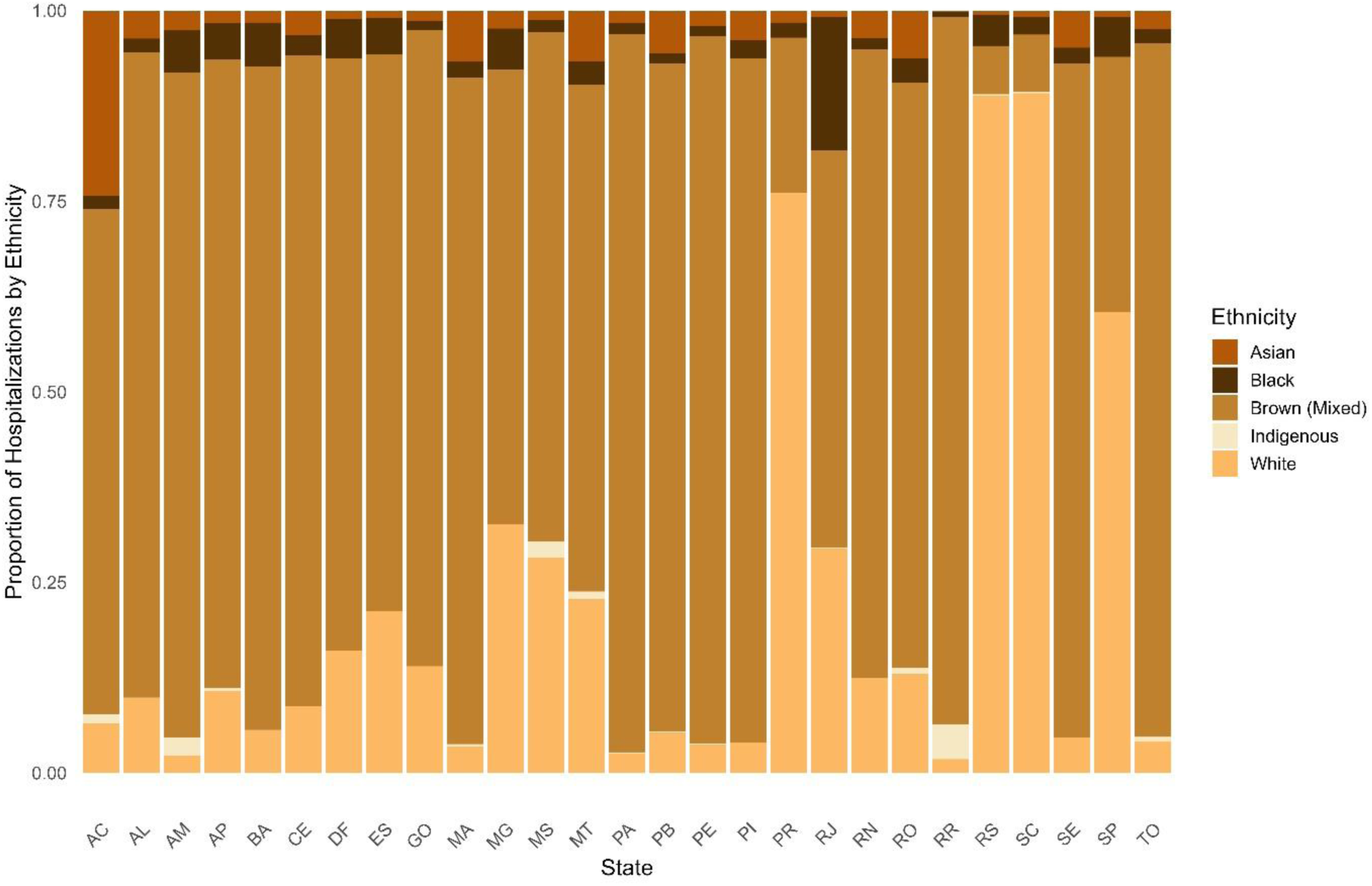
Distribution of fracture-related hospitalizations by ethnicity across Brazilian states, 2015–2024. Stacked bar chart displaying the proportional distribution of SIH/SUS-recorded hospitalizations for each ethnicity category within each Brazilian state.

Relative to fracture type across ethnicities, fractures of other limb bones accounted for the highest proportion overall, particularly among Indigenous individuals (73.1%), who also exhibited the largest share of hospitalizations due to multiple region/body fractures (11.6%). White individuals showed the highest proportion of femur fractures (20.6%), while Black individuals had the greatest number of hospitalizations for fractures of the skull and facial bones (Figure 7).

**Figure 7.**
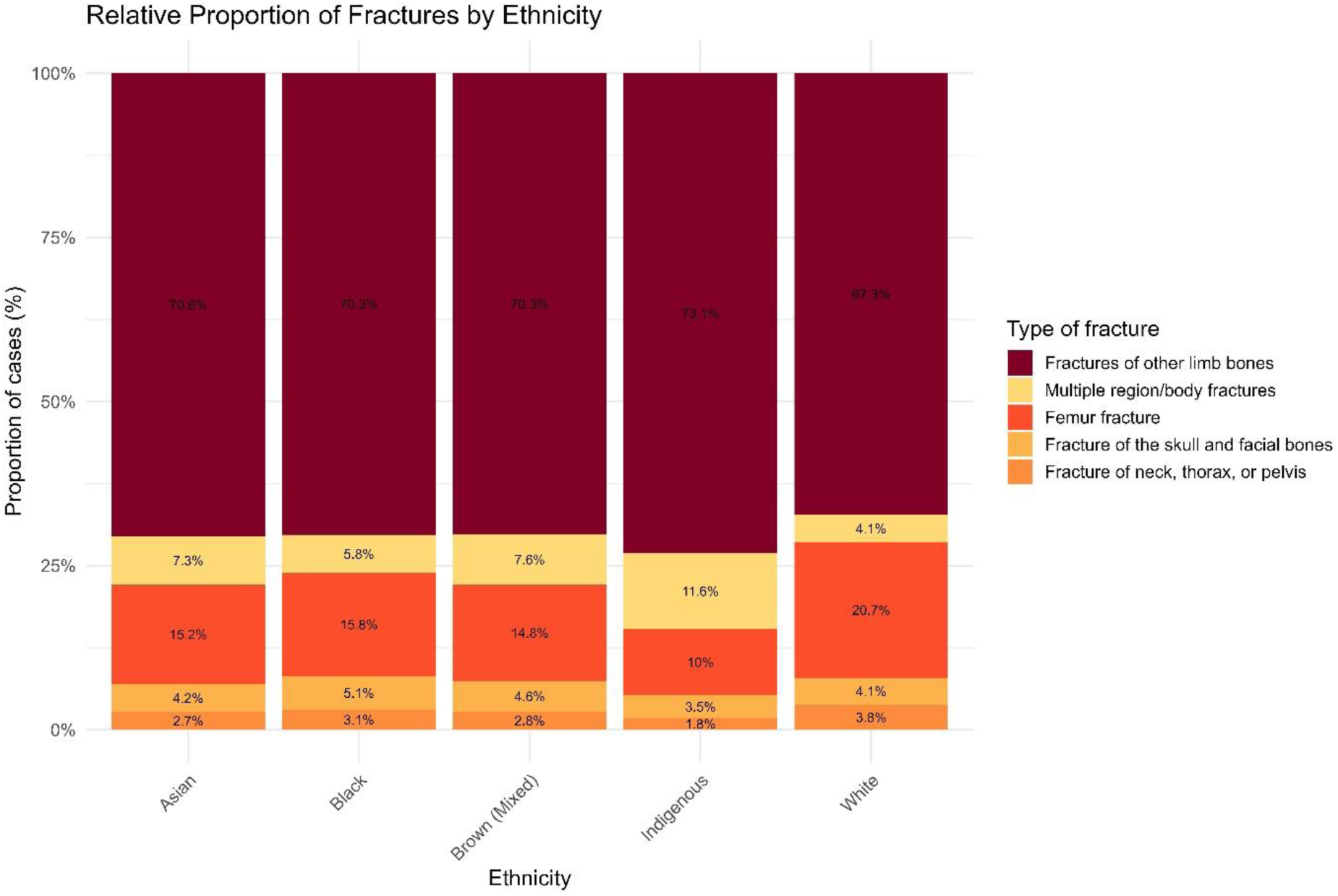
Relative distribution of fracture types across ethnic groups in Brazil, 2015–2024. Stacked bar chart showing the proportional distribution of SIH/SUS-recorded fracture-related hospitalizations within each ethnic group across five fracture categories.

## Discussion

In the period analyzed in this study (2015 – 2024), Brazil exhibited a steady increase in hospitalizations due to bone fractures. This upward trend parallels the global patterns reported by the Global Burden of Disease Study 2019, which estimated 178 million new fractures worldwide in 2019, a 33.4% increase since 1990, along with 455 million prevalent cases and 25.8 million years lived with disability (YLDs) (Vos *et al*. 2020). The estimated annual percent change (APC) further corroborates this sustained rise in fracture-related hospitalizations in Brazil. Brazil mirrors the global scenario, with a progressive rise in absolute fracture burden despite potential stabilization in age-standardized rates. In both contexts, the rise in absolute numbers can be related to the population aging and growth (Ferreira *et al*. 2023). It is important to note that the global increase (33.4%) reported by the *GBD 2019* represents a cumulative growth between 1990 and 2019, whereas the Brazilian APC reflects an average yearly increase between 2015 and 2024. Although they describe different time scales and statistical metrics, both indicate a consistent upward trend in fractures burden, with the Brazilian APC suggesting a more accelerated recent growth compared to the long-term global trajectory. The most pronounced increases in femoral and other limb fractures underscore the growing impact of osteoporotic fragility fractures among older adults, compounded by injuries related to falls and traffic accidents (Vasconcelos *et al*. 2020; Lima, Salles and Silva 2022). As observed globally, these trends highlight the urgent need for preventive strategies targeting bone health, fall prevention, and trauma reduction, particularly in an aging population.

The analysis of sex-related fracture patterns reveals that the occurrence of fractures is not independent of sex. Men experienced nearly twice the number of hospitalizations between 2014 and 2024 compared to women. This is supported by a studied with Pernambuco’ patients, where a 2:1 ratio was observed between hospitalizations for femur fractures and for fractures of other limb bones in men compared with women, reinforcing the vulnerability of this group (Maciel *et al*. 2012). Futhermore, this disparity is widely documented in the literature, which consistently reports a higher incidence of traumatic and high-energy fractures in males due to greater exposure to motor-vehicle collisions, occupational hazards, and risk-taking behaviors (Almigdad et al. 2022a, 2022b; Huang et al. 2023).

Sex is a key determinant in the epidemiology of fractures, shaping both their frequency and the mechanisms that lead to injury. The analysis of hospitalizations between men and women by fracture type revealed a marked epidemiological divergence in fracture-related hospitalizations, a pattern widely documented in the literature (Tonini and Nazário 2021; Moreira, Nissan and Gontijo 2025). Among young and middle-aged men (20–59 years), the high burden of fractures of other limb bones, along with the predominance of skull, facial, and thoracic/pelvic injuries is consistent with high-energy trauma mechanisms (Arruda *et al*. 2009; Sindeaux 2024). In contrast, the pattern observed in women is substantially different; hospitalizations rise only at older ages, particularly among those aged 80 years or older, with femur fractures becoming the dominant injury type. The literature supports this finding, reporting a higher prevalence of proximal femur fractures in women and highlighting the biomechanical vulnerability associated with postmenopausal osteoporosis, which predisposes older women to low-energy fractures, most commonly resulting from ground-level falls increases and contributes to fracture risk (Vasconcelos *et al*. 2020; Castro Mendes *et al*. 2023; Ferreira *et al*. 2023). In a retrospective study encompassing 397,585 hospitalizations due to femur fractures within the Brazilian Unified Health System (SUS), the predominance of female cases was also observed, with an average annual incidence of 213.83 per 100 000 older adults (Macedo *et al*. 2019). These contrasting etiologies, high-energy trauma in men versus bone fragility in older women, underscore the need for targeted injury-prevention and public-health strategies that account not only for sex differences but also for life-course stage and the predominant mechanisms of injury within each group.

The analysis of years of life lost (YLL) further reinforces the divergent mechanisms underlying fracture-related outcomes in men and women. Femur fractures accounted for a large proportion of total YLL and yet were characterized by relatively low mean YLL per death, reflecting mortality concentrated later in life, which is corroborated with number of hospitalizations by femur fracture in the 80 year and over (Johnell and Kanis 2006; Cooper *et al*. 2011). Interestingly, Silva et al. (2024) showed that male sex and age are risk factors for increased one-year mortality following femur surgery. This higher mortality has been attributed to multifactorial causes, including smoking and alcohol consumption, combined with severe comorbidities, particularly pulmonary and cardiovascular diseases, which accounted for 36.4% and 18.2% of deaths, respectively (Silva *et al*. 2024). In contrast to the YLL of femur fractures, the fracture types more common in younger men, such as skull, facial, and thoracic/pelvic injuries, contributed proportionally less to total YLL but exhibited markedly high mean YLL values, indicating premature mortality consistent with high-energy trauma mechanisms discussed (Haagsma *et al*. 2016; Vos *et al*. 2020). The comparison of mean YLL across sexes showed that males consistently experienced greater years of life lost for every fracture category, aligning with the higher lethality of early-life traumatic injuries in this group. Taken together, these findings confirm that while women bear the burden of fracture-related mortality at advanced ages due to skeletal fragility, men disproportionately experience premature death driven by high-impact trauma (Kingma 1994). These sex-specific trajectories highlight the need for complementary public-health approaches, including traffic and occupational safety policies broadly targeting men, alongside osteoporosis management and fall-prevention strategies directed toward older women (Gillespie *et al*. 2012).

In our dataset, the Northeast region included states with some of the lowest numbers of total fracture hospitalizations, such as Amazonas, Acre, Roraima, and Amapá, however also included a state with the highest count of hospitalization, which was Rondônia. This pattern reflects the overall distribution of all fracture types, which encompasses a wide range of injuries with different mechanisms, severities, and healthcare pathways. In contrast to this data, in a study conducted by Modesto et al. (2022), covering the period from 2008 to 2021 and focusing exclusively on femoral fractures, the region with the highest number of hospitalizations was the Southeast, followed by the Northeast, with males being the most frequently hospitalized sex (Modesto, Ribeiro and Pereira 2022). Our analysis also pointed out Piauí as one of the states with the higher number of hospitalizations. Peterle et al. (2020) reported that Piauí registered 6 480 652 hospitalizations for femur fracture in the population over 60 years old in Brazil, between 2008 and 2018, most of which occurred in women aged 80 years or older (PETERLE *et al*. 2020), similar to our findings about gender profile of femur-related hospitalizations. However, in contrast to our data, Soares reported that the higher total number of hospitalizations, between 2011 and 2016, occurred in the Southeast region, with this region also showing the highest occurrence of femoral fracture–related admissions in both females and males (Soares *et al*. 2020). It is important to note that studies frequently focus on femoral fractures because they represent one of the most clinically severe and resource-intensive type of musculoskeletal injury. Hip and femur fractures are strongly associated with high mortality, long hospital stays, the need for surgical intervention, and substantial costs to the health system (Veronese and Maggi 2018; Vasconcelos *et al*. 2020). However, this femur-specific focus introduces important limitations. While femur fractures provide valuable information about severe orthopedic injuries and elderly health, they offer only a partial view of the national fracture profile, meaning that broader and more comprehensive datasets are still needed to fully understand the epidemiology of fractures across different age groups, mechanisms of injury, and regions.

With respect to the ethnic distribution of hospitalized patients, findings from previous literature contrast with the results of the present analysis. In a study by Siqueira et al (2005), White patients accounted for the largest share of hospitalizations, followed by Black patients and then individuals categorized as mixed-race (Siqueira, Facchini and Hallal 2005). These differences may relate to underlying biological and demographic factors, as Black individuals tend to achieve a higher peak bone mass compared with White individuals, a characteristic often associated with lower fracture risk and reduced prevalence of osteoporosis (Seeman 1998; Bryant *et al*. 2003). When compared with other groups, Indigenous individuals were the most affected by fractures involving multiple body regions and by injuries classified as “other body regions”. Black and brown individuals showed a higher occurrence of skull and facial fractures, whereas white individuals were more frequently affected by femoral fractures. The association between skull and facial fractures and Black and brown populations reflects socioeconomic rather than biological determinants. These injuries, typically resulting from motorcycle accidents or interpersonal violence, are linked to high-energy trauma (Conceição *et al*. 2013; Maia *et al*. 2022). According to the 2022 Census, 72.9% of residents of favelas and urban informal settlements self-identify as Black or brown (IBGE, 2022). Regarding Indigenous individuals, the 2022 Census reveals that their population grew by approximately 89% between 2010 and 2022, and that over half (about 54%) of Indigenous people now live in urban areas, including outside Indigenous territories (Instituto Brasileiro de Geografia e Estatística (IBGE) 2023). These figures suggest that the higher occurrence of multiple fractures and fractures in other body regions among Black/brown and Indigenous populations may reflect broader socioeconomic vulnerability, urbanization processes and violence and differential exposure to high-energy trauma mechanisms.

## CONCLUSION

This nationwide analysis reveals a sustained increase in fracture-related hospitalizations in Brazil, alongside marked disparities across sex, age, fracture type, region, and ethnicity, underscoring the complex and multifactorial nature of fracture burden in the country. Men were disproportionately affected by high-energy trauma, resulting in greater premature mortality, while women experienced a sharp concentration of femur fractures at older ages, reflecting the growing influence of population aging and skeletal fragility. Regional and ethnic inequalities further highlighted social and structural vulnerabilities that shape exposure to trauma and access to care. By providing a comprehensive and disaggregated overview of national trends, this study offers critical evidence to guide the development of targeted public-health policies, ranging from traffic and occupational injury prevention to fall-prevention and osteoporosis management, and to strengthen trauma care systems in the most vulnerable populations. Such epidemiological monitoring is essential for informed decision-making, resource allocation, and the design of effective strategies aimed at reducing fractures and improving outcomes across Brazil.

### Author contributions

DCB and PPA contributed to the conceptualization of the study. PPA collected and analyzed the data and wrote the manuscript. DCB reviewed the manuscript and supervised the study. All authors approved the final version.

### Supplementary data

Supplementary data is available at IJE online.

### Conflict of interest

None declared.

## Funding

This study was funded by the National Council for Scientific and Technological Development – CNPq and Brazilian Ministry of Health through the DECIT/SECTICS/MS/CNPq call (Process No. 445067/2023-3), by the *Programa Jovem Cientista do Nosso Estado* (Process No. E-26/201.403/2022 – fellowship), and by the Brazilian National Program of Genomics and Precision Health – Genomas Brasil (DECIT/SECTICS/MS/CNPq; Process No. 444206/2023-0).

### Data availability

The data used in this study are publicly available from the Hospital Information System of the Brazilian Unified Health System (SIH/SUS), provided by the Brazilian Ministry of Health through DATASUS (Brazilian Ministry of Health, 2024), and can be accessed at http://tabnet.datasus.gov.br/cgi/deftohtm.exe?sih/cnv/nruf.de.

### Use of artificial intelligence tools

Artificial intelligence (AI) tools were not used in the preparation of this manuscript.

## Data Availability

All data produced in the present work are contained in the manuscript

## Notes

### Competing Interest Statement

The authors have declared no competing interest.

### Author Declarations

The datas used in the study is already de-identified at the database source itself.

## References

1. Albergaria B-H, Zerbini CAF, Szejnfeld VL et al. An updated hip fracture incidence rate for Brazil: the Brazilian Validation Osteoporosis Study (BRAVOS). Arch Osteoporos 2022;17:90.

2. Almigdad A, Mustafa A, Alazaydeh S et al. Bone Fracture Patterns and Distributions according to Trauma Energy. Adv Orthop 2022;2022:1–12.

3. Arruda LRP, Silva MA de C, Malerba FG et al. Fraturas expostas: estudo epidemiológico e prospectivo. Acta Ortop Bras 2009;17:326–30.

4. Brazilian Ministry of Health. Hospital Information System of the Unified Health System (SIH/SUS). DATASUS. Brasília: Ministry of Health; 2024. Accessed August, 2025.

5. Bryant RJ, Wastney ME, Martin BR et al. Racial Differences in Bone Turnover and Calcium Metabolism in Adolescent Females. J Clin Endocrinol Metab 2003;88:1043–7.

6. Castro Mendes M, De Oliveira Alemães JP, Malheiros Monteiro B et al. Fatores de risco de fratura de fêmur em idosos: uma revisão bibliográfica. Brazilian Journal of Implantology and Health Sciences 2023;5:6094–103.

7. Cauley JA. Public Health Impact of Osteoporosis. J Gerontol A Biol Sci Med Sci 2013;68:1243–51.

8. Conceição LD, Lund RG, Nascimento GG et al. Non-white people have a greater risk for maxillofacial trauma: findings from a 24-month retrospective study in Brazil. Braz J Oral Sci 2013;12:313–8.

9. Cooper C, Cole ZA, Holroyd CR et al. Secular trends in the incidence of hip and other osteoporotic fractures. Osteoporosis International 2011;22:1277–88.

10. Ferreira GRON, Chagas T de N das C e, Gonçalves LHT et al. Fall-Related Hospitalizations in Elderly People: Temporal Trend and Spatial Distribution in Brazil. Geriatrics 2023;8:30.

11. Gillespie LD, Robertson MC, Gillespie WJ et al. Interventions for preventing falls in older people living in the community. Cochrane Database of Systematic Reviews 2012;2021, DOI: 10.1002/14651858.CD007146.pub3.

12. Haagsma JA, Graetz N, Bolliger I et al. The global burden of injury: incidence, mortality, disability-adjusted life years and time trends from the Global Burden of Disease study 2013. Injury Prevention 2016;22:3–18.

13. Huang B, Wang Y, Wang H et al. Epidemiology and the economic burden of traumatic fractures in China: A population-based study. Front Endocrinol (Lausanne) 2023;**Volume** 14**-2023.**

14. Instituto Brasileiro de Geografia e Estatística (IBGE). Censo Demográfico 2022: Resultados Gerais. Rio de Janeiro, 2023.

15. Johnell O, Kanis JA. An estimate of the worldwide prevalence and disability associated with osteoporotic fractures. Osteoporosis International 2006;17:1726–33.

16. Kingma J. The Young Male Peak in Different Categories of Trauma Victims. Percept Mot Skills 1994;79:920–2.

17. Lima JA, Salles LP, Silva MAM da. Perfil Epidemiológico de Idosos Internados por Fratura de Fêmur no Brasil. Revista de Saúde 2022;13:59–65.

18. Macedo GG, Gomes Teixeira TR, Ganem G et al. Fraturas do fêmur em idosos: um problema de saúde pública no Brasil. Revista Eletrônica Acervo Científico 2019;6:e1112.

19. Maciel SSSV, Maciel WV, Lima Neto AJ de et al. Internação hospitalar por fraturas de fêmur e outros ossos dos membros em residentes de Pernambuco. Rev Assoc Med Rio Grande Do Sul 2012;56:213–9.

20. Maia SÉ da S, Cardoso LIS, Moreira TCAraújo et al. Análise epidemiológica das fraturas dos ossos da face em um hospital público no nordeste do Brasil . Brazilian Journal of Oral and Maxillofacial Surgery 2022;22:6–12.

21. Mensor L, Rosim M, Marasco G et al. Avaliação de custos associados a fraturas por fragilidade no Sistema Único de Saúde (SUS) e no Sistema de Saúde Suplementar (SSS) no Brasil. Jornal Brasileiro de Economia da Saúde 2021;13:288–99.

22. Modesto WHGC, Ribeiro EA, Pereira F de A. Internações hospitalares por fratura de fêmur no Brasil e suas regiões: série temporal de 2008 a 2021. Research, Society and Development 2022;11:e100111436119.

23. Moreira MVS, Nissan IN, Gontijo T de C. Estudo comparativo sobre fratura de fêmur em idosos: uma análise dos anos 2020 e 2023 e suas relações com a pandemia de COVID-19. PECIBES 2025;11:22–7.

24. Peterle Vcu, Geber Junior Jc, Darwin Junior W, Et Al. Indicators of Morbidity and Mortality by Femur Fractures in Older People: A Decade-Long Study In Brazilian Hospitals. Acta Ortop Bras 2020;28:142–8.

25. Sarmento JP da F, Silva FR da, Aranda IB et al. Custos com a internação hospitalar por fraturas de fêmur em idosos, no Brasil, entre 2016 e 2020. Research, Society and Development 2022;11:e214111739153.

26. Seeman E. Growth in Bone Mass and Size—Are Racial and Gender Differences in Bone Mineral Density More Apparent than Real? J Clin Endocrinol Metab 1998;83:1414–9.

27. Silva ABF, Silva MCP da, Lima GK et al. Hip fracture in Pará State, Brazil: officially recorded mortality and comorbidities in the elderly population. Retrospective cohort study. Rev Panamazonica Saude 2024;15, DOI: 10.5123/S2176-6223202401381.

28. Sindeaux LMES. Epidemiologia do trauma de face em um hospital referência no estado do Ceará. 2024.

29. Siqueira FV, Facchini LA, Hallal PC. The burden of fractures in Brazil: A population-based study. Bone 2005;37:261–6.

30. Soares EB, Lira NET de, Silva MWLA da et al. Fraturas de Fêmur – panorama das taxas de morbimortalidade e incidência entre as regiões brasileiras. Tópicos Em Ciências Da Saúde - Volume 18. Editora Poisson, 2020.

31. Tonini SF, Nazário NO. Perfil Epidemiológico De Fratura Proximal De Fêmur Em Idosos Atendidos Em Um Hospital Geral Da Grande Florianópolis E Sua Associação Com Sexo E Idade. Arquivos Catarinenses de Medicina 2021;50:23–35.

32. Vasconcelos PAB de, Rocha A de J, Fonseca RJ de S et al. Femoral fractures in the elderly in Brasil - incidence, lethality, and costs (2008-2018). Rev Assoc Med Bras 2020;66:1702–6.

33. Veronese N, Maggi S. Epidemiology and social costs of hip fracture. Injury 2018;49:1458–60.

34. Vos T, Lim SS, Abbafati C et al. Global burden of 369 diseases and injuries in 204 countries and territories, 1990–2019: a systematic analysis for the Global Burden of Disease Study 2019. The Lancet 2020;396:1204–22.

35. Wu A-M, Bisignano C, James SL et al. Global, regional, and national burden of bone fractures in 204 countries and territories, 1990–2019: a systematic analysis from the Global Burden of Disease Study 2019. Lancet Healthy Longev 2021;2:e580–92.

