## Supplementary material for "Trends in Fracture-Related Hospitalizations and Mortality in Brazil, 2015–2024 Bone Fracture Trends in Brazil": Tables

**Hospitalizations and mortality due to bone fractures in Brazil, 2015–2024: an epidemiological perspective**

**TABLES**

**Table 1. Annual Percent Change (APC) in Fracture-Related Hospitalizations, 2014–2024**

| **Fracture type** | **APC** | **95% CI** | **p-value** |
| --- | --- | --- | --- |
| Femur fracture | 4.31 | 3.76–4.85% | p < 0.001 |
| Fracture of the neck, thorax, or pelvis | 4.47 | 3.58–5.36% | p < 0.001 |
| Fracture of the skull and facial bones | 1.61 | –0.28–3.52 | p = 0.134 |
| Fractures of other limb bones | 4.51 | 3.65–5.39% | p < 0.001 |
| Multiple region/body fractures | 6.13 | 5.29–6.97% | p < 0.001 |

CI: Confidence Interval

|  | **Hospitalizations** | | **Hospitalizations per Fracture (%)** | | **Odds Ratio** | **95% CI** | **Deaths** |  | **Fatality (%)** |  |
| --- | --- | --- | --- | --- | --- | --- | --- | --- | --- | --- |
| **Fracture type** | **Female** | **Male** | **Female** | **Male** |  |  | **Female** | **Male** | **Female** | **Male** |
| Femur fracture | 340,944 | 514,981 | 16.5 | 11.3 | 1.55 | 1.54-1.56 | 23,379 | 1,4587 | 6.86 | 2.83 |
| Fracture of neck. thorax. or pelvis | 56,201 | 154,350 | 2.7 | 3.4 | 0.80 | 0.79-0.81 | 1,261 | 4,051 | 2.24 | 2.62 |
| Fracture of the skull and facial bones | 56,994 | 256,554 | 2.8 | 5.6 | 0.48 | 0.50-0.51 | 292 | 1,684 | 0.51 | 0.66 |
| Fractures of other limb bones | 1,477,107 | 3,336,181 | 71.6 | 73.4 | 0.92 | 0.91-0.92 | 2,317 | 4,633 | 0.16 | 0.14 |
| Multiple region/body fractures | 131,020 | 284,274 | 6.4 | 6.3 | 1.02 | 1.01-1.02 | 4,235 | 4,042 | 3.23 | 1.42 |

**Table 2. Sex-specific Hospitalization Rates, Mortality, and Odds Ratios for Different Fracture Types in Brazil, 2015–2024.**

**Table 3. Mean Years of Life Lost (YLL) and Proportional YLL Distribution by Fracture Type and Sex.**

| **Age** | **YLL** | | | |
| --- | --- | --- | --- | --- |
|  | **Mean (years)** | | **Proportion (%)** | |
|  | **Female** | **Male** | **Female** | **Male** |
| Femur fracture | 10.0 | 14.0 | 25.8 | 22.3 |
| Fracture of the neck, thorax, or pelvis | 23.0 | 27.0 | 2.97 | 11.3 |
| Fracture of the skull and facial bones | 32.0 | 34.0 | 0.92 | 5.62 |
| Fractures of other limb bones | 19.0 | 28.0 | 4.6 | 13.4 |
| Multiple region/body fractures | 11.0 | 19.0 | 5.26 | 7.84 |
